# Global Burden, Temporal Trends, and Health Inequalities of Early-Onset Group B Streptococcus Infection in Neonates, 1990–2021: A Systematic Analysis of the Global Burden of Disease Study 2021

**DOI:** 10.64898/2026.04.02.26350022

**Authors:** Qiang Wen, Xiu Wang, Yanjing Wu, Yayun Jiang, Zhihong Xu

## Abstract

**Objectives:** Group B Streptococcus (GBS) is a leading cause of neonatal mortality worldwide. However, the global burden of early-onset GBS disease (EOD-GBS) has not been fully elucidated. We aimed to describe the geographical distribution and epidemiological characteristics of the EOD-GBS burden, and analyze its association with socio-economic development and universal health coverage.

**Methods:** We used data from the Global Burden of Disease Study 2021 and the Universal Health Coverage Service Coverage Index (UHC-SCI) to calculate estimated annual percentage changes (EAPCs) of EOD-GBS mortality. Sex differences were analyzed using the conservative overlap assessment. The geographical distribution of EOD-GBS clinical presentations and mortality was mapped. Health inequality analysis was conducted to evaluate the relationship between the sociodemographic index (SDI), UHC-SCI, and EOD-GBS burden.

**Results:** Global EOD-GBS mortality decreased by nearly 50% from 1990 (693.41 per 100,000) to 2021 (348.80 per 100,000). However, the decline was not uniform: the most significant decrease occurred in high-middle SDI regions (EAPC: −7.17%), and the slowest in low SDI regions (EAPC: −2.23%). Male neonates accounted for the most EOD-GBS deaths, particularly in high SDI regions. Lower respiratory infections were common in Asia and Oceania; meningitis was more prominent in Europe. Inequality analysis revealed a phenomenon of “absolute convergence but relative differentiation”: as social development and universal health coverage improves, the absolute mortality gap between countries narrowed, but relative burden concentrated increasingly among the poorest populations.

**Conclusions:** The global burden of EOD-GBS has decreased substantially, but there are marked differences among countries. Continued socioeconomic development and expanded universal health coverage are critical to further reduce neonatal mortality.

## Introduction

Group B Streptococcus (GBS) remains a leading cause of neonatal infection worldwide. Invasive GBS (iGBS) typically manifests as severe sepsis, meningitis, or lower respiratory tract infections (LRIs), with a high mortality rate [1]. iGBS is categorized based on neonatal onset timing as early-onset disease (EOD-GBS; 0–6 days) or late-onset disease (LOD-GBS; 7–89 days) [2]. EOD-GBS is primarily acquired through vertical transmission, either through ascending amniotic infection or aspiration during passage through the birth canal. LOD-GBS is typically associated with horizontal transmission from environmental sources such as hospitals. Neonates with EOD-GBS exhibit more severe clinical presentations, with a mortality risk of 5–27% [2].

The global EOD-GBS burden exhibits distinct geographical heterogeneity. The estimated global incidence is 0.41 per 1,000 live births. However, regional disparities are striking, with incidence ranging from lows of 0.20 in South Asia and 0.32 in other parts of Asia, to 0.72 in Africa, and peaking at 1.47 in the Caribbean [2]. Mortality rates mirror these variations. The fatality rate is approximately 5% in developed countries, rising significantly in less-developed regions and countries and reaching 27.0% in Africa [2]. The United Nations has called for enhanced effective prevention measures targeting the major causes of neonatal mortality to further reduce global neonatal mortality and disease burden. However, comprehensive assessments of the global EOD-GBS disease burden remain limited [2–4], necessitating multidimensional evaluations to guide and refine prevention strategies for neonatal EOD-GBS.

The Global Burden of Disease (GBD) 2021 database of GBS pathogens provides an opportunity to analyze and assess the global burden of neonatal EOD-GBS disease, offering mortality and disability-adjusted life-year (DALY) metrics for 204 countries and regions [5]. The incidence and prognosis of EOD-GBS are linked to the quality of maternal and neonatal healthcare, such as prenatal screening coverage, intrapartum antibiotic prophylaxis (IAP) implementation [6], and skilled birth attendant availability [4]. We introduced the reproductive, maternal, newborn, and child health (RMNCH) subindex (indicator 3.8.1) from the Universal Health Coverage Service Coverage Index (UHC-SCI) defined by the World Health Organization [7]. This index reflects a country’s coverage of maternal and child health services within essential health services. By integrating the GBD database with national essential health service coverage data, this study aimed to map the geographic distribution and temporal trends of the EOD-GBS burden and explore the association between disease burden and socioeconomic development using the sociodemographic index (SDI).

## 2. Materials and Methods

### 2.1 Data sources and case definitions

We used data from the GBD Study 2021, published in 2024. The GBD Study 2021 provides a comprehensive assessment of the incidence, mortality, and DALYs for 371 diseases and injuries across 204 countries and territories between 1990 and 2021 [5]. We extracted data on GBS infections in early neonates. The primary metrics analyzed were mortality rates and DALYs, stratified by sex, location, and year. As the burden of disease in neonates aged 0–6 days is predominantly driven by premature mortality (years of life lost) and acknowledging the intrinsic collinearity between DALY and mortality rates in this age group, our statistical analysis focused on mortality metrics to avoid redundancy.

Ethical approval was waived by the Ethics Committee of Deyang People’s Hospital because of the use of de-identified publicly available data. This study adheres to the Guidelines for Accurate and Transparent Health Estimates Reporting (GATHER) [8].

### 2.2 Statistical analysis

#### 2.2.1 Temporal trends

To quantify the temporal trends in the disease burden of EOD-GBS between 1990 and 2021, we calculated the estimated annual percentage change (EAPC). The EAPC values and their 95% confidence intervals (CIs) were calculated using a log-linear regression model. An increasing trend was indicated if the lower limit of the 95% CI was positive; a decreasing trend was indicated if the upper limit was negative.

#### 2.2.2 Assessment of sex disparities

To rigorously evaluate whether EOD-GBS mortality differs significantly by sex, we adopted the “conservative overlap assessment” based on the non-overlapping of uncertainty intervals (UIs). We calculated a difference metric defined as follows:

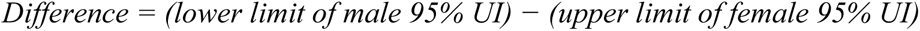

A difference of >0 indicates a significantly higher mortality rate in males than in females (no overlap). A difference of <0 suggests no significant difference under this conservative threshold. Values closer to zero indicated borderline significance.

#### 2.2.3 Socio-economic and health coverage stratification

We used the SDI and UHC-SCI to assess the relationship between disease burden, national socioeconomic development levels, and health service coverage. The SDI is a composite indicator of development status derived from lag-distributed income per capita, average educational attainment for those aged ≥15 years, and the total fertility rate for those under 25 [5]. Countries were categorized into five quintiles based on 2021 SDI scores: low (<0.46), low-middle (0.46–0.61), middle (0.61–0.69), high-middle (0.69–0.81), and high (0.81–1.00). To evaluate the impact of maternal and child health services, we extracted the RMNCH sub-index from the WHO UHC Service Coverage Index (indicator 3.8.1). This geometric mean of the four tracer indicators reflects the coverage of essential services across the continuum of care from pregnancy to childhood, reported on a unitless scale from 0 to 100 [7].

#### 2.2.4 Inequality analysis

We employed the slope index of inequality (SII) and concentration index to quantify cross-country inequalities in EOD-GBS mortality [9]. The SII (absolute inequality) was calculated by regressing mortality rates against the relative ranks of countries stratified by SDI or UHC-SCI-RMNCH. It represents the absolute difference in predicted values between the most and least advantaged populations. The concentration index (relative inequality) is derived from the numerical integration of the area under the Lorenz curve, which models the cumulative proportion of deaths against the cumulative population ranked by SDI or UHC metrics [10]. For both indices, negative values indicate that the disease burden was disproportionately concentrated in countries with lower socioeconomic or health coverage levels. The magnitude of this value reflects the degree of inequality.

#### 2.2.5 Software

All statistical analyses and visualizations were performed using R software (version 4.3.3) and RStudio (version 2023.06.2+561). Statistical significance was defined as a two-sided P-value of <0.05.

## 3. Results

### 3.1 Global and regional trends in mortality and DALYs (1990–2021)

Between 1990 and 2021, the global burden of neonatal EOD-GBS significantly decreased. The total number of EOD-GBS-related deaths decreased by approximately 50%, from 17,606 in 1990 to 8,550 in 2021 (Table 1). Correspondingly, the global mortality rate declined from 693.41 to 348.80 per 100,000 population, representing an EAPC of −2.37%. Regarding sex disparities, a distinct male predominance was observed in mortality burden; in 2021, the mortality rate for male neonates was 404.58 per 100,000, higher than that for female neonates (289.04 per 100,000). When stratified by SDI, EOD-GBS-related deaths were predominantly concentrated in the low-middle SDI (3,008 cases) and low-SDI (4,530 cases) regions, exceeding those in the high-middle SDI (69 cases) and high-SDI (14 cases) regions. Regarding EAPCs, high-middle SDI regions exhibited the largest absolute reduction (−7.17%) and low-SDI regions showed the smallest absolute reduction (−2.23%).

**Table 1.**
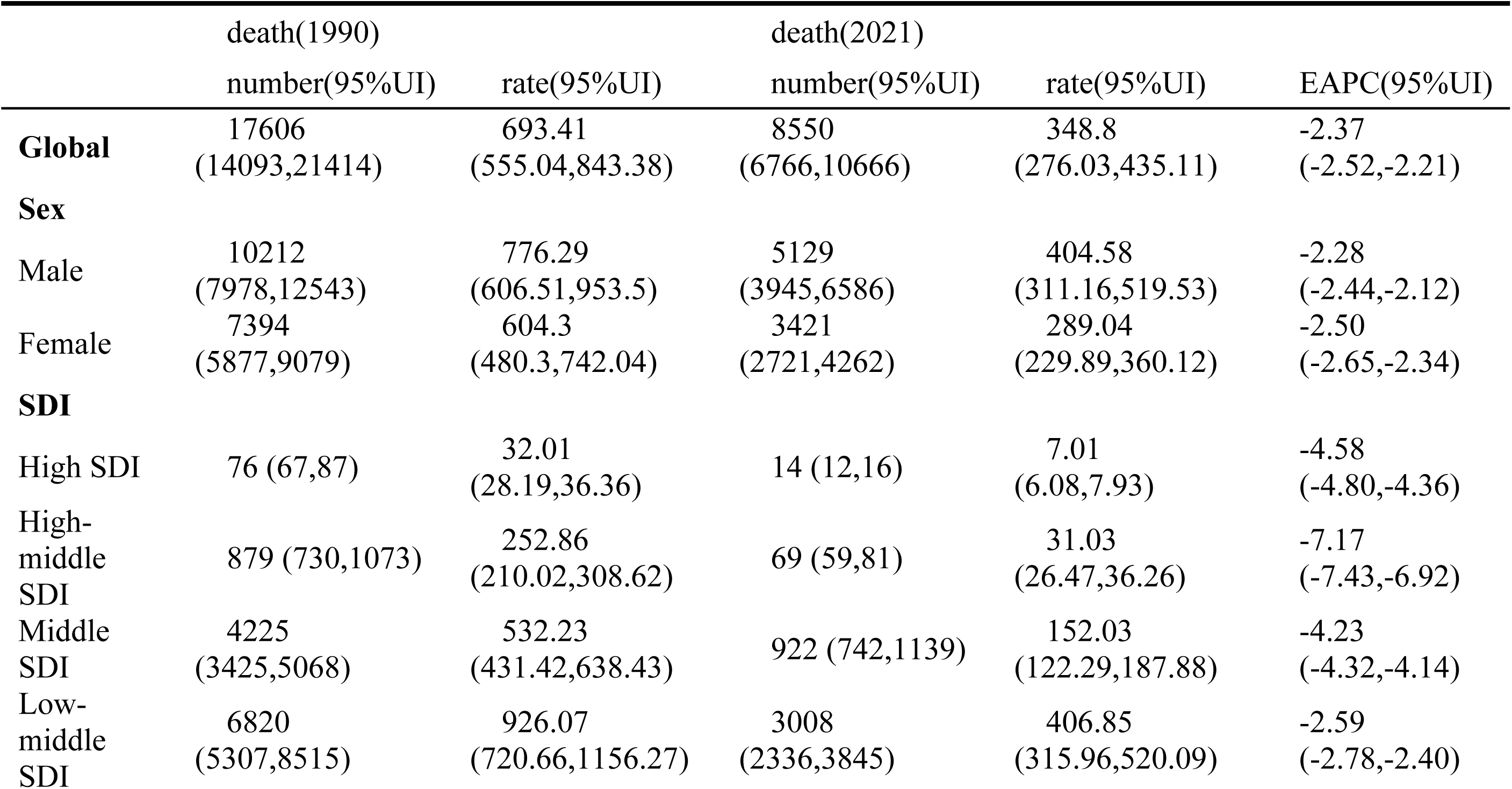
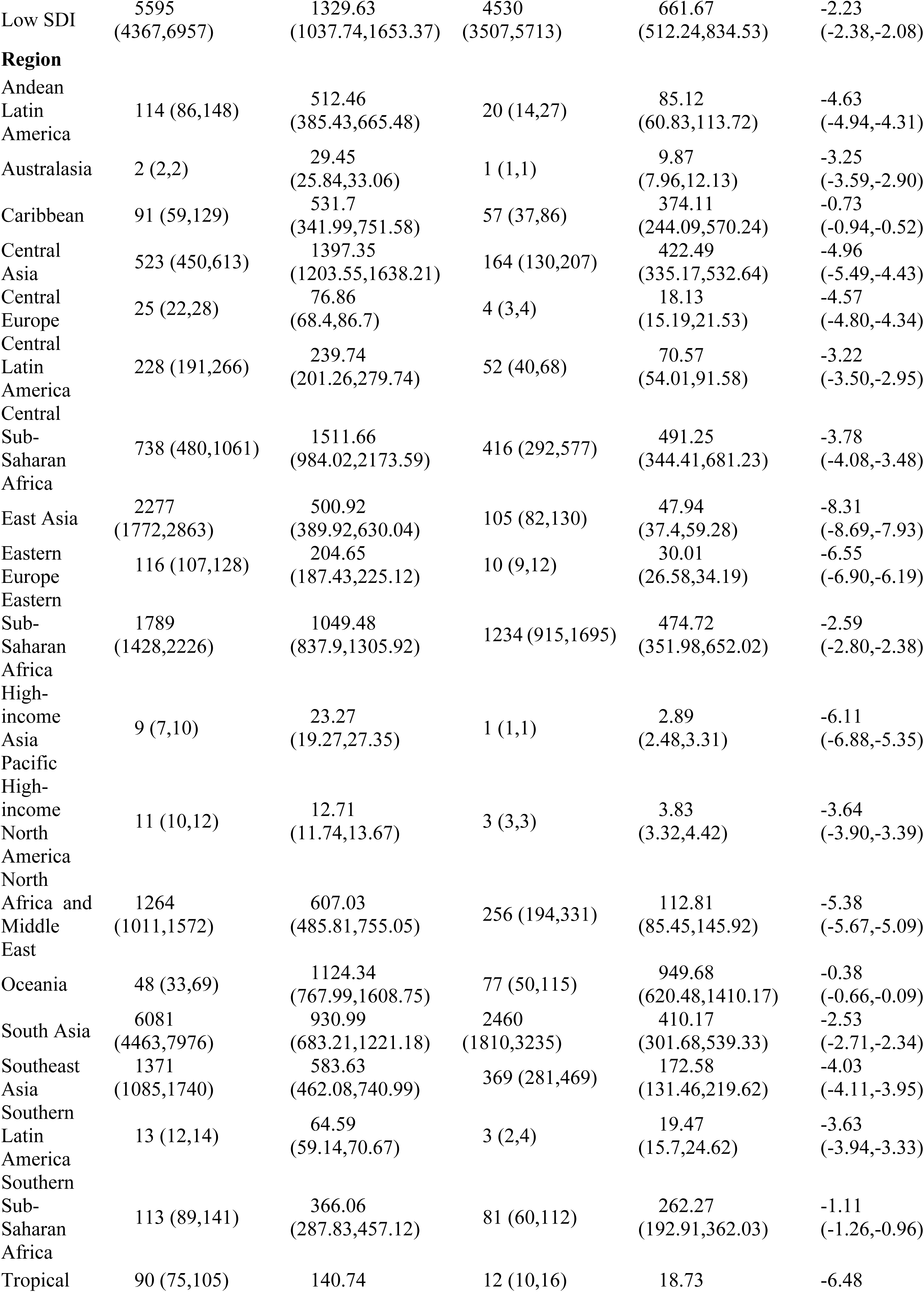
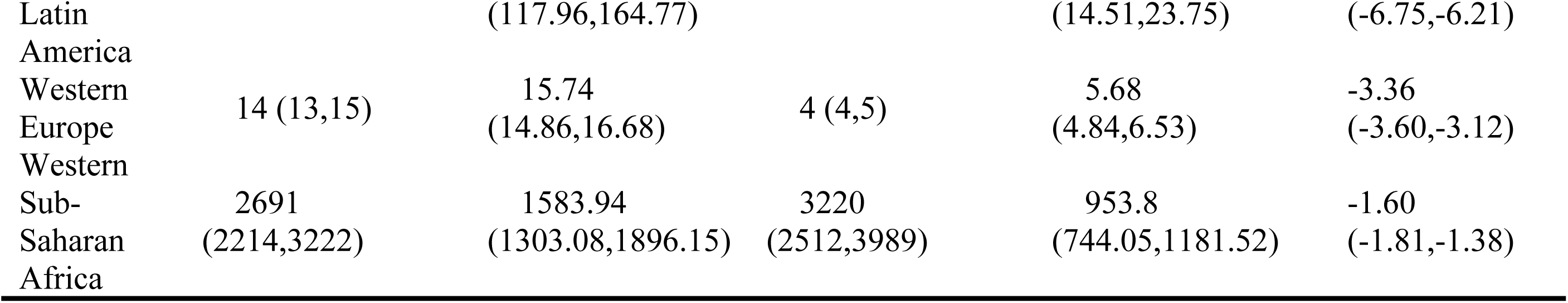
Global and regional EOD-GBS Death cases, mortality rates, and Estimated Annual Percentage Change for 1990 and 2021.

Geographically, EOD-GBS burden distribution exhibited marked heterogeneity across the 21 GBD regions. Western sub-Saharan Africa had the highest number of deaths (3,220), followed by South Asia (2,460). The lowest figures were observed in high-income Asia Pacific and Australasia (1 each). Western sub-Saharan Africa and Oceania had the highest mortality rates of 953.8 and 949.68 deaths per 100,000 live births, respectively. The high-income Asia-Pacific and high-income North American regions had the lowest rates of 2.89 and 3.83 per 100,000, respectively. East Asia demonstrated the most significant improvement over the study period (EAPC: −8.31%), whereas Oceania showed the least (EAPC: −0.38%). DALYs trends mirrored those of mortality; in 2021, the global burden amounted to 769,284 DALYs, with a rate of 31,383.76 per 100,000 people. Similar to the mortality indicators, the DALY rates were higher in males (31,383.76) than in females (26,007.36) and followed identical temporal and geographical patterns (see Additional file 1).

### 3.2 Geographical distribution, country-level burden, and temporal trends

The EOD-GBS burden demonstrated remarkable intercountry variability (Figure 1A; Additional file 2). In 2021, Nigeria recorded the highest neonatal mortality rate (1,215.97 per 100,000; 95% UI 934.05–1,557.77) and Andorra reported the lowest (0.30; 95% UI 0.05–0.66). Regionally, Western sub-Saharan Africa bore the heaviest burden (953.80 per 100,000), followed by Oceania (949.68 per 100,000). Conversely, high-income Asia-Pacific (2.89) and North America (3.83) had the lowest burdens (see Additional file 3).

**Figure 1.**
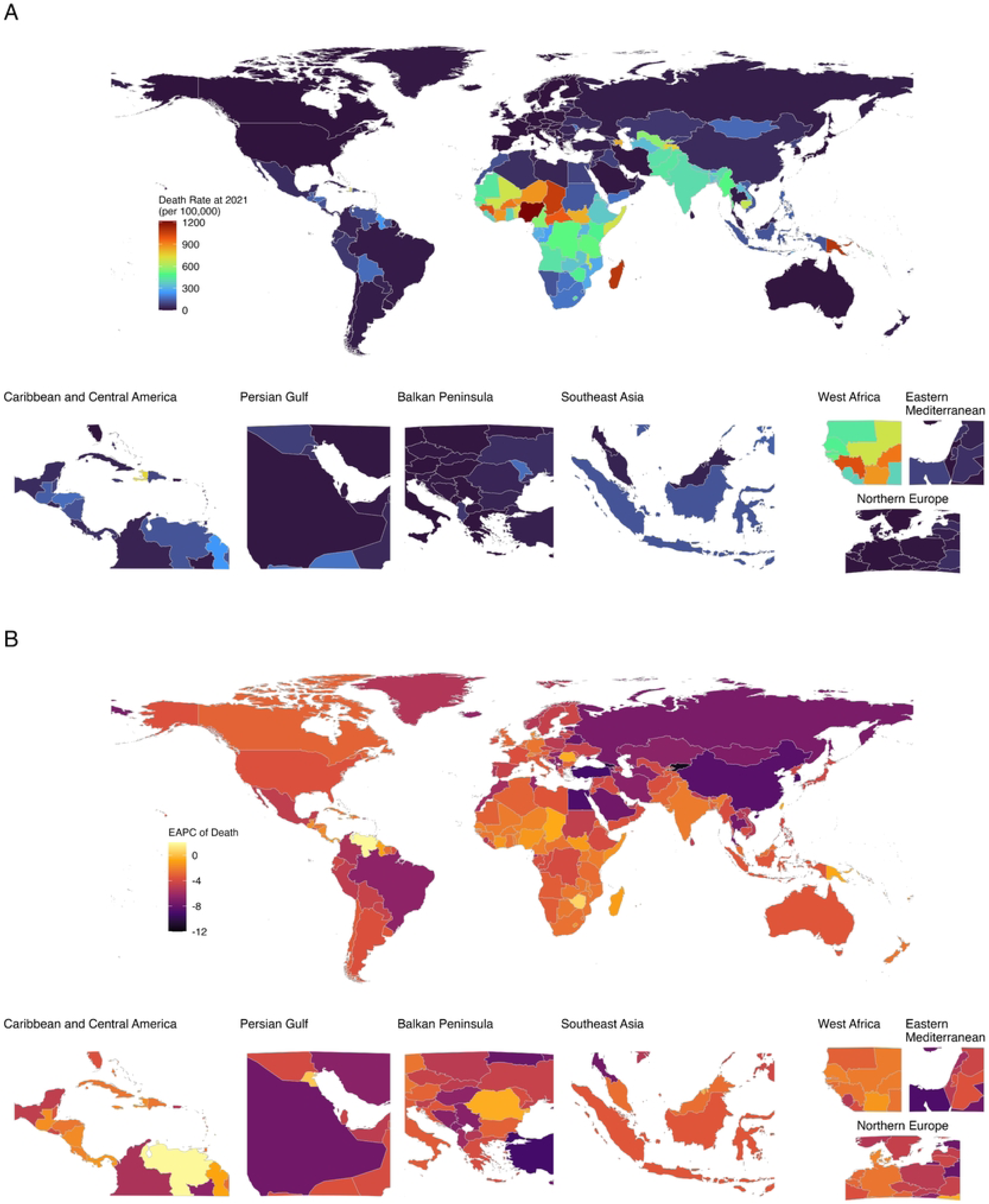
EOD-GBS neonatal mortality rates (2021) and EAPC (1990–2021) across 204 countries. A. EOD-GBS neonatal mortality rate in 2021; B. EAPC of EOD-GBS neonatal mortality rates between 1990 and 2021.

Between 1990 and 2021, Kyrgyzstan (−11.43%) and Georgia (−10.41%) achieved the fastest annual declines (EAPCs) in mortality (Figure 1B; Additional file 4). However, three countries exhibited increasing mortality rates: Venezuela (1.81%), Dominica (1.01%), and Zimbabwe (0.67%). At the regional level, East Asia (−8.31%) and Eastern Europe (−6.55%) experienced the most rapid improvements, while Oceania (−0.38%) and the Caribbean (−0.73%) showed the slowest progress. All 21 geographical regions exhibited declining mortality rates over time (see Additional file 5).

Between 1990 and 2021, EOD-GBS mortality rates significantly declined globally and across all five SDI quintiles (Figure 2), though temporal trends varied by development level. High and high-middle SDI regions exhibited a biphasic decline, with rapid early reductions followed by later deceleration. Conversely, low and low-middle SDI regions exhibited more monotonic, near-linear decreases. Male mortality consistently exceeded female mortality across all SDI regions. Interestingly, the magnitude of this sex disparity was more pronounced in the high and high-middle SDI regions than in lower-income settings, with the gap widening over the latter half of the study period.

**Figure 2.**
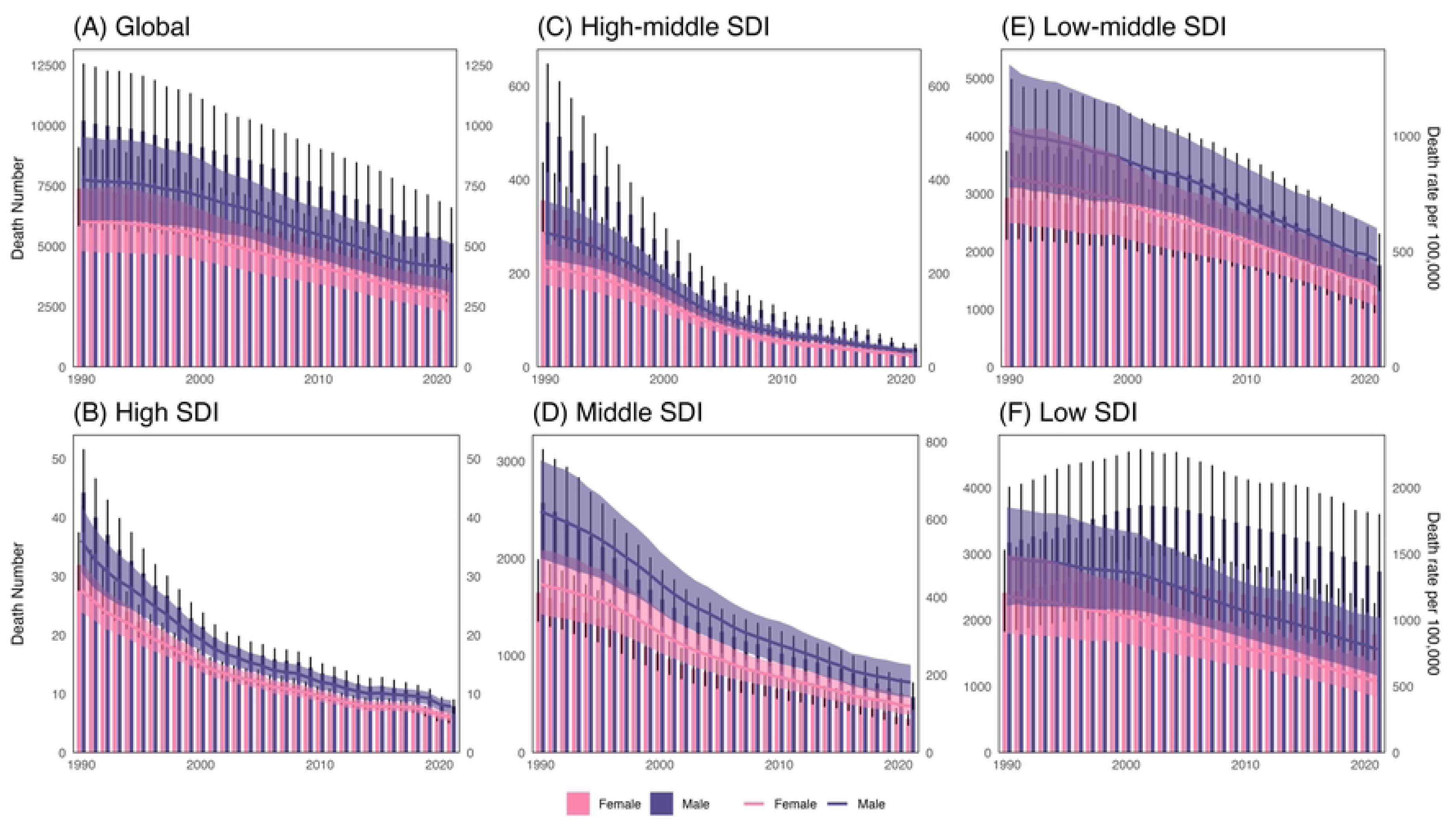
Trends in EOD-GBS mortality and death counts globally and across different SDI regions, 1990–2021.

### 3.3 Sex disparities in mortality

In 2021, 177 of the 204 countries (86.8%) exhibited male dominance in EOD-GBS mortality rates (Figure 3A; Additional file 6). We further explored the association between sex disparities and socioeconomic development (Figure 3B); the “conservative overlap assessment” revealed a positive correlation between the magnitude of sex disparities and SDI (R = 0.351, p<0.05). This indicates that as the SDI increases, the significance distinguishability of the sex gap widens. Specifically, nine countries within the high-to-middle SDI bracket showed significantly higher male mortality than female mortality (overlap value >0). Contrastingly, in low-and middle-SDI countries, while the mean male mortality was higher than that for females, the difference was rarely significant owing to wider uncertainty intervals.

**Figure 3.**
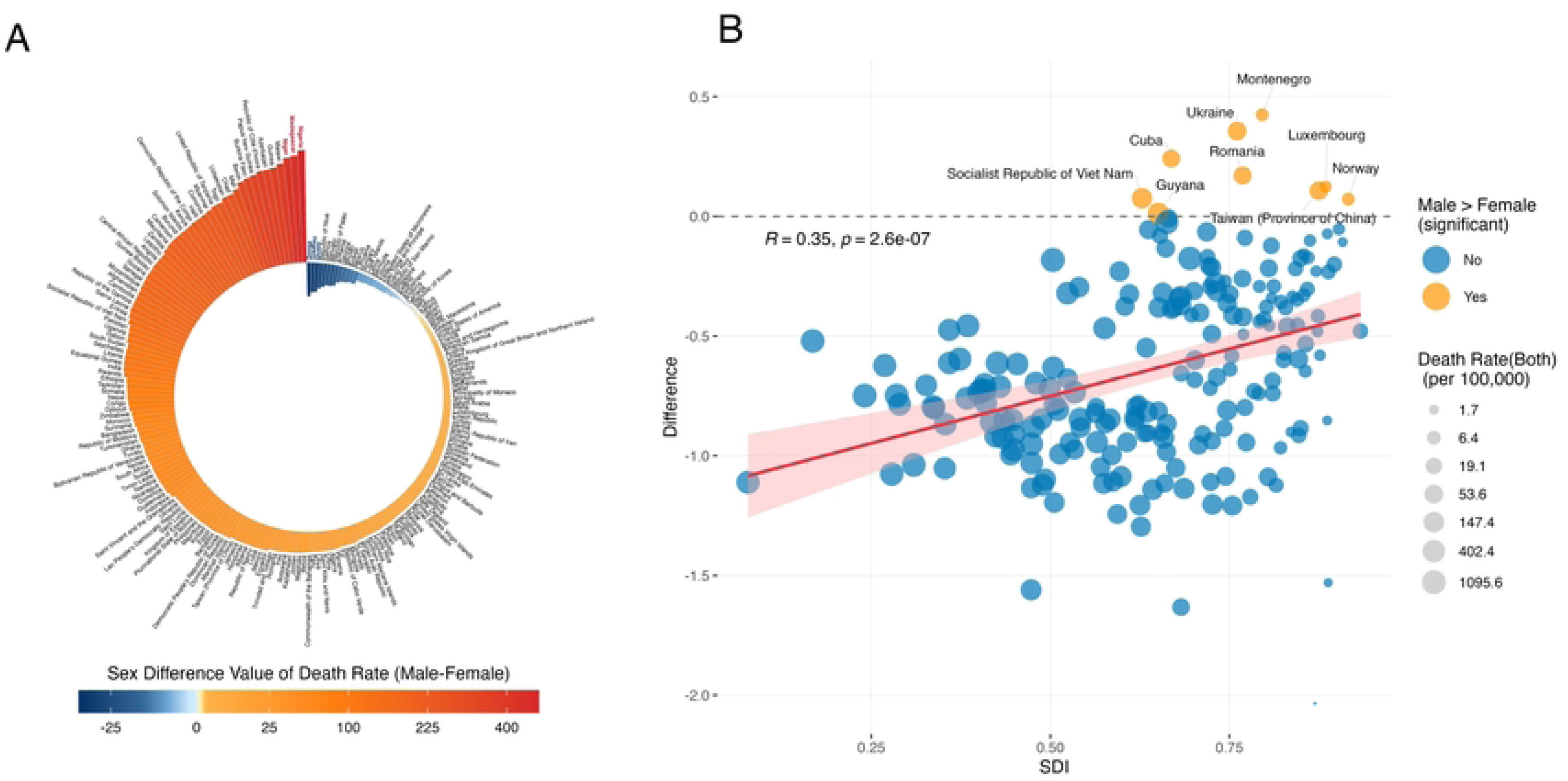
Analysis of Sex Difference in Neonatal Mortality Rates from EOD-GBS. A. Map showing differences in male and female infant mortality rates across 204 countries in 2021; B. Correlation analysis between neonatal sex difference and SDI across 204 countries.

### 3.4 Syndromic distribution: the role of LRI

In 2021, LRIs accounted for 81.8% of global EOD-GBS deaths and was the dominant clinical presentation. However, this rate varied across geographical regions (Figure 4). Oceania consistently reported the highest LRI rates, reaching 98.3% in 2021, followed by Central, East, South, and Southeast Asia (range: 89.7–95.9%). Contrastingly, Western Europe reported the lowest historical LRI rate (43.5% in 2021). Between 1990 and 2021, the overall ranking of LRI rates across the 21 regions remained largely stable. Significant changes were noted in Central Europe (from 66.7% to 80.5%) and Southern Latin America (from 55.6% to 70.3%). At the country level, Switzerland (22.6%) had the lowest LRI rate, whereas Uzbekistan (98.7%) had the highest (see Additional file 7). No significant correlation was found between the SDI and proportional contribution of LRIs (see Additional file 8).

**Figure 4.**
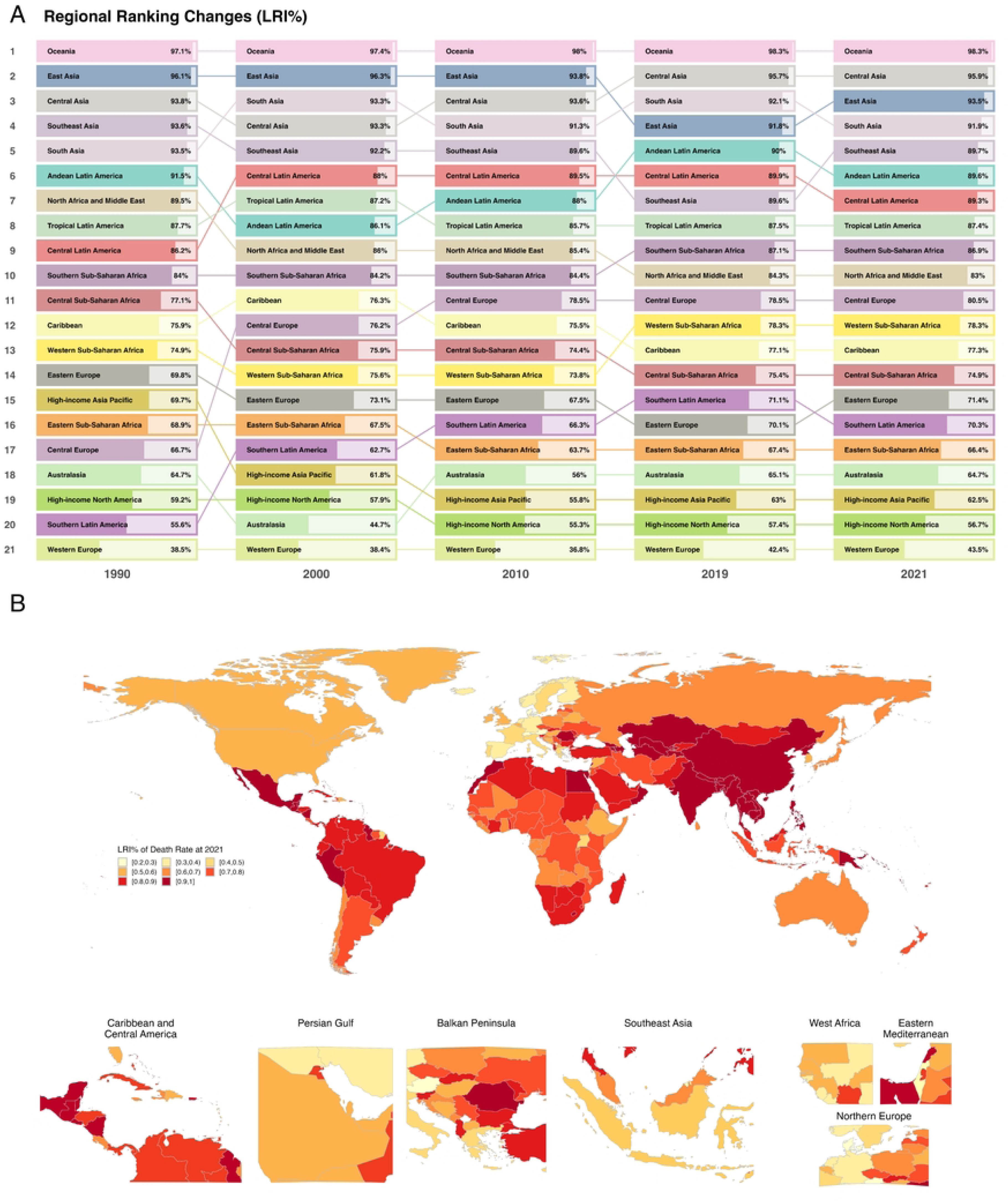
Trends in the proportion of lethal EOD-GBS clinical manifestations and the distribution of LRI proportions. A. Changes in the proportion of LRI within neonatal mortality rates for EOD-GBS across 21 regions between 1990 and 2021. B. Distribution of LRI proportions within EOD-GBS mortality rates across 204 countries in 2021.

### 3.5 Health inequality analysis and association with UHC service coverage

Health inequality analysis of EOD-GBS mortality using the SDI and UHC-SCI-RMNCH index revealed a complex dynamic of “absolute convergence” but “relative divergence” (Figure 5, Additional file 9). The absolute inequality (SII) indicated a significant narrowing of the absolute gap in mortality between the highest and lowest SDI countries, dropping from 1,330 deaths per 100,000 in 1990 to 559 in 2021. A similar convergence was observed for the UHC-SCI-RMNCH index (1,218 in 2000 and 598 in 2021), indicating that the absolute burden in the most disadvantaged countries is decreasing. Conversely, the concentration index for SDI became more negative (from −0.48 in 1990 to −0.58 in 2021), with the difference primarily originating from high and middle SDI regions. This indicates that the remaining disease burden was increasingly concentrated in low-SDI regions. The UHC-SCI-RMNCH-based concentration index also showed increasing relative inequality (−0.52 in 2000 to −0.61 in 2021), but the difference was more evenly distributed across all SDI regions.

**Figure 5.**
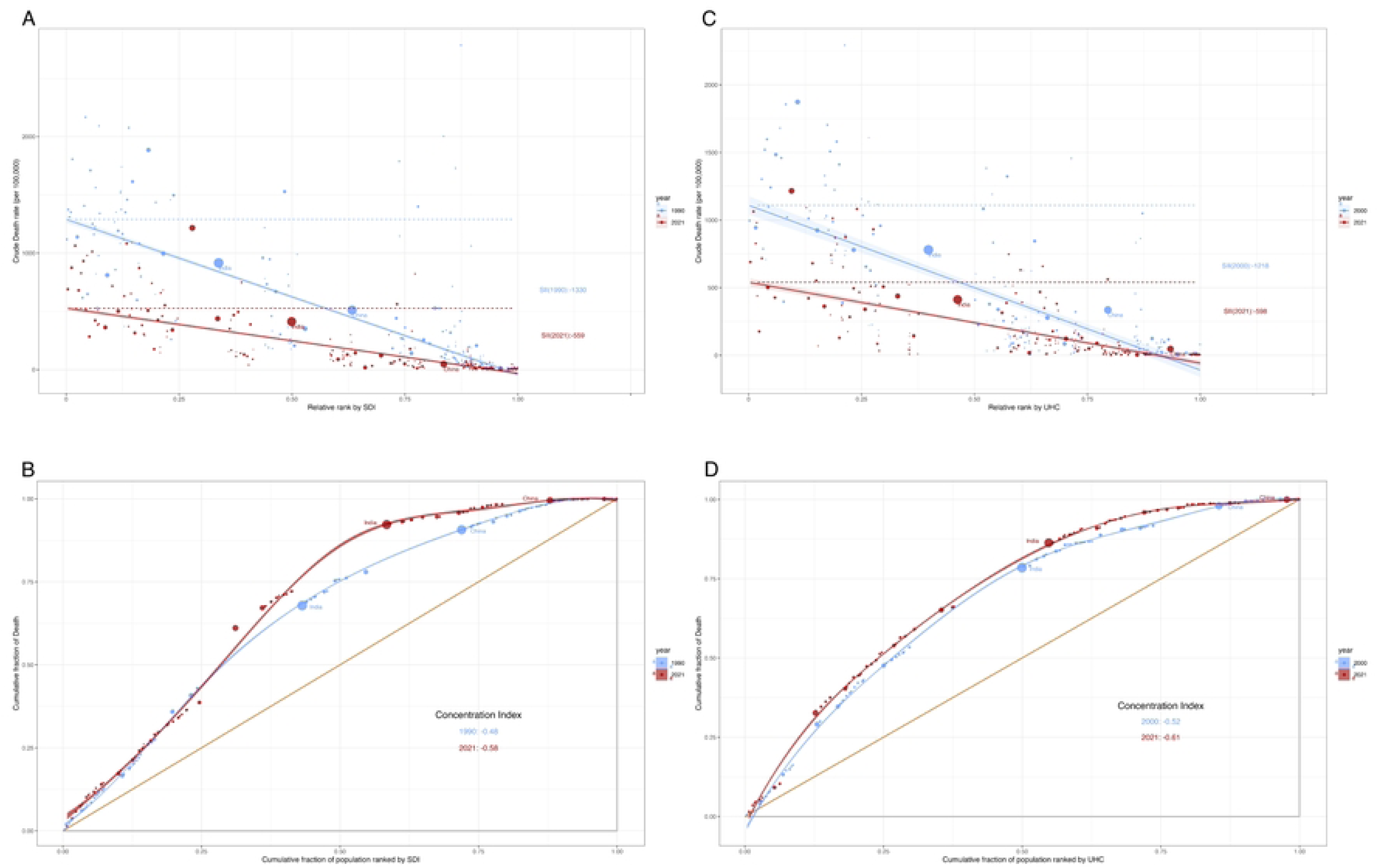
Regression and concentration curves for health inequalities in SDI and UHC-SCI-RMNCH versus global EOD-GSB mortality. Regression curves (A) and concentration curves (B) for health inequalities in SDI versus global EOD-GSB mortality in 1990 and 2021. Regression curves (C) and concentration curves (D) for health inequalities in UHC-SCI-RMNCH versus global EOD-GSB mortality in 2000 and 2021.

Regression analysis revealed a robust inverse association between the UHC-SCI-RMNCH index and EOD-GBS mortality. Each 1-unit increase in the UHC index was associated with a reduction in EOD-GBS mortality by 12.8–19.7 per 100,000 (p<0.001), although the protective effect attenuated over time (coefficient change: −19.7 in 2000 to −12.8 in 2021) (Figure 6). After adjustment for SDI, the effect size of the UHC index was reduced by approximately 52% (coefficient reduced to −5.17 in 2021), indicating that about half of the observed benefits reflect broader socioeconomic development, with the remainder attributable to RMNCH-specific health service coverage.

**Figure 6.**
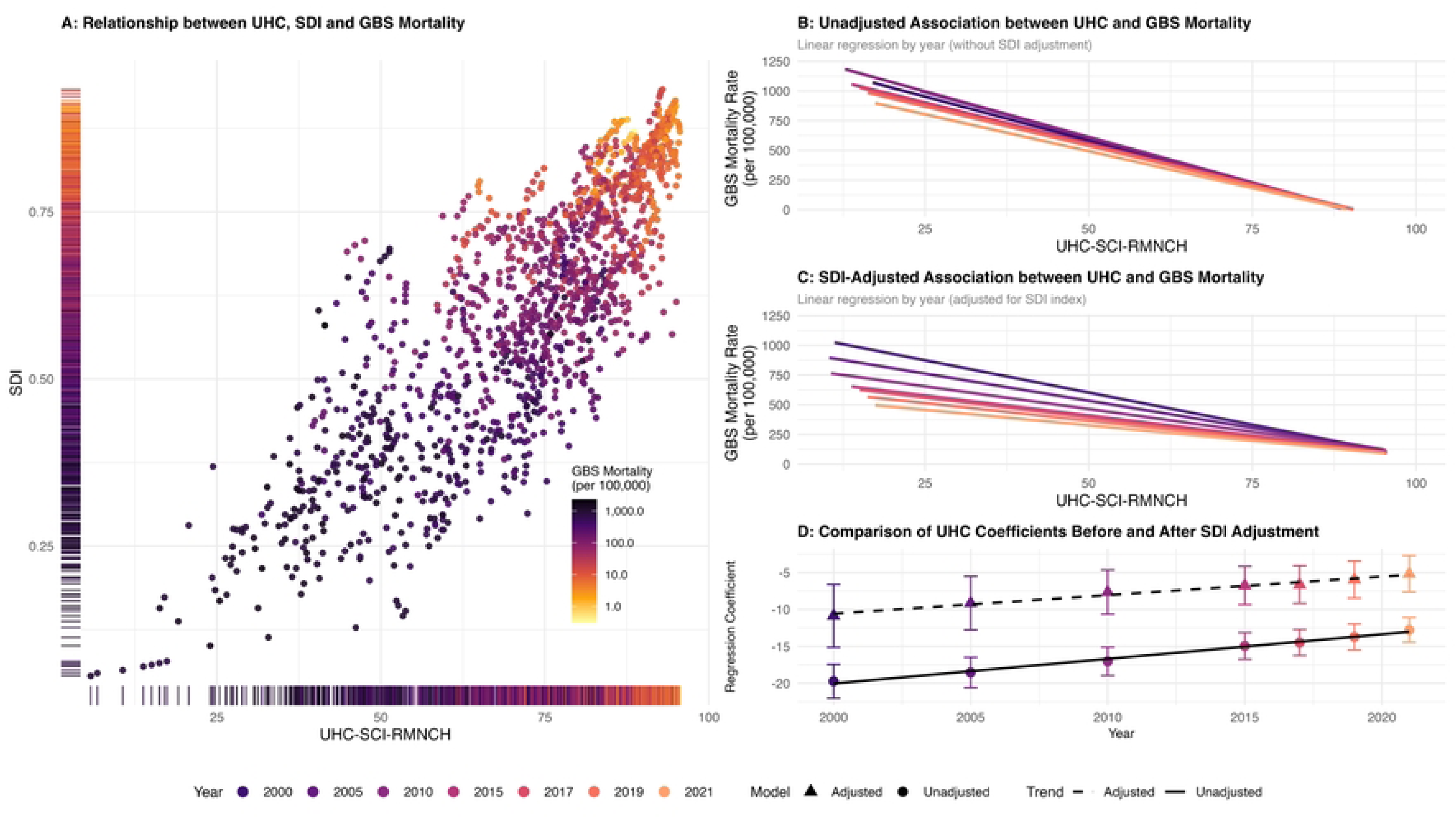
Correlation between UHC-SCI-RMNCH Index and EOD-GBS Mortality. A. Relationship between GBS mortality and UHC/SDI across countries; B. Linear regression analysis of UHC and GBS mortality; C. Linear regression analysis of UHC and GBS mortality adjusted by SDI index; D. Comparison of regression coefficients over time before and after model adjustment.

## 4. Discussion

In response to the United Nations’ mandate to reduce preventable neonatal mortality, this study provides a comprehensive assessment of the global burden of EOD-GBS between 1990 and 2021 based on the GBD Study 2021 data. Our findings highlight a substantial reduction in the global burden, with both mortality rates and death counts halving over the past 32 years. This is closely linked to efforts made by the global effort in preventing and controlling EOD-GBS. For instance, the revised guidelines for preventing early-onset GBS issued by the United States in 2002 [11], along with risk-based screening strategies implemented in countries such as the United Kingdom and Sweden [12], have played a crucial role in reducing neonatal deaths caused by EOD-GBS. The stark contrast in the EAPC between high-middle SDI regions (−7.17%) and lower SDI regions underscores the critical role of socioeconomic development in EOD-GBS control. High-resource settings successfully leveraged economic advantages to minimize impacts, whereas low-resource settings achieved more moderate gains, maintaining a consistent annual decline in EOD-GBS mortality rates.

A persistent male predominance in EOD-GBS mortality was observed globally, aligning with the “male disadvantage” hypothesis in neonatal sepsis [13, 14]. Similar findings were reported in the UK and France, with both showing a higher incidence of EOD-GBS in male infants than in female infants [15]. Further, in the Netherlands, GBS meningitis mortality was significantly higher among boys aged <3 months than among girls [16]. Indeed, sex significantly influences the outcome of multiple infectious diseases; this may be because females exhibit stronger humoral and cellular immune responses to infection or antigenic stimulation than males, which aids in defending against and eliminating pathogens [17]. Interestingly, our analysis revealed that the magnitude of sex differences was positively correlated with SDI. We hypothesize that this phenomenon is driven by “ascertainment bias” rather than biology alone. In high-SDI regions, timely access to sophisticated neonatal care and precise diagnostics ensures that nearly all cases are identified, thereby revealing the “true” biological gap in survival outcomes. Conversely, in low SDI regions, the limitations of healthcare infrastructure and incomplete case reporting may obscure these subtle sex differences.

The global pattern of the EOD-GBS burden exhibits extreme polarization, with a 4,000-fold disparity in mortality rates between the highest (Nigeria) and lowest (Andorra) burden countries in 2021. Among the 21 regions, Africa exhibited significantly higher mortality rates than other regions, with extreme disparities exceeding 300-fold, reflecting the social determinants of health. Three decades of sustained social development have driven declines in EOD-GBS neonatal mortality in most countries; however, the rising trends in Venezuela, Dominica, and Zimbabwe serve as cautionary warnings. In these countries, political instability and economic crises have likely precipitated the collapse of health systems, leading to a drain of medical personnel and shortage of medical supplies, resulting in a sharp increase in maternal and infant mortality rates. This reversal of progress underscores the fact that sustained political and economic stability is a crucial prerequisite for ensuring a continued reduction in the EOD-GBS burden.

Neonatal EOD-GBS may lead to severe clinical manifestations such as meningitis and LRIs. Globally, meningitis accounts for a lower proportion than LRIs, primarily because meningitis typically presents later and often manifests as LOD-GBS [18]. A nationwide Japanese study found that meningitis cases accounted for 26.9% of the EOD-GBS incidence, lower than the 36.5% incidence in LOD [19]. Our study highlights geographical variations in the clinical presentation of fatal EOD-GBS; LRI was dominant in the Asia-Pacific region, while meningitis exceeded 40% in Western Europe and North America, matching the LRI prevalence. We propose two possible explanations for this variation. First, surveillance bias plays a critical role; in resource-poor settings, meningitis reporting is not mandatory, and the lethal nature of neonatal meningitis often results in infant death before microbiological confirmation, complicating etiological assessment and leading to underestimation of GBS meningitis incidence [18]. Conversely, high-income countries possess robust surveillance systems capable of capturing complex cases. Second, microbiological heterogeneity is a contributing factor. Serogroup III is the predominant serogroup in GBS meningitis [18] and is frequently associated with the highly virulent Clonal Complex 17 carrying the meningitis-prone hvgA gene [20]. Serogroup III is predominant in the UK (60%) [15], but less prevalent in certain regions of South America and Asia [21]. These geographical variations in serogroup distribution partly explain the regional differences in the distribution of EOD-GBS as a cause of neonatal mortality.

Leveraging the UHC-SCI-RMNCH index and SDI, we uncovered a complex dynamic of “absolute convergence” but “relative divergence” in health inequality across countries regarding the EOD-GBS burden between 1990 and 2021. While the absolute gap in mortality between the most and least developed nations has narrowed, the relative concentration of the burden on the poor has intensified. The relative inequality based on SDI primarily intensified due to “leapfrog” progress in high and middle SDI regions, reflecting the critical role of socioeconomic development in achieving the near-zero mortality target for neonatal EOD-GBD. Conversely, the widening inequality based on UHC was relatively evenly distributed across the UHC index spectrum, indicating diminishing health benefits from the UHC index growth over time.

Our regression analysis further indicated a diminishing protective effect of UHC over time (diminishing marginal returns), which was heavily contingent on broader socioeconomic development. The independent effect of UHC (adjusted for SDI) decreased by approximately 52%; this implies that ∼50% of UHC’s health gains were fundamentally rooted in the enabling environment created by a nation’s overall socioeconomic development level. This explains why high UHC nearly eliminated GBS mortality in high SDI countries (e.g., Luxembourg at 3.6 cases per 100,000), while in low SDI countries, even with improved UHC, the “efficiency” of translating coverage into health outcomes remained low, exacerbating relative inequalities. Overall, the EOD-GBS disease burden exhibits a persistent and severe “pro-poor” phenomenon. Only by combining universal health services with socioeconomic development, pursuing both tracks simultaneously, can inequality be gradually eliminated.

This study had several limitations. First, the widespread absence of high-quality vital registration data in low-SDI regions, coupled with the high concentration of EOD-GBS disease burden, may have led to an underestimation of the findings. Second, reliance on the GBD Study 2021 database, whose data sources and modeling assumptions have limitations, affects the reliability of the conclusions [22]. Third, the UHC-SCI-RMNCH data were available for a limited number of years, which may have affected the precision of our long-term trend analysis. Finally, the conclusions drawn from the GBD Study 2021 data may be subject to the potential impacts of the COVID-19 pandemic period.

## 5. Conclusions

The global landscape of neonatal EOD-GBS has witnessed a promising 50% reduction in mortality over the past three decades; however, this overall reduction in disease burden masks persistent pro-poor inequalities. Improvements have been primarily driven by socioeconomic development and the expansion of basic health coverage, yet the “low-hanging fruit” of foundational interventions appears to have been harvested. Our findings indicate diminishing marginal returns from UHC, particularly in lower-SDI settings, suggesting that the existing paradigm of solely expanding service coverage has reached its limit. Furthermore, the relative widening of inequalities and the emergence of sex disparities in developed regions underscore the critical role of robust surveillance and health system quality. To achieve the sustainable development goal of ending preventable neonatal deaths, future strategies must transcend traditional biomedical approaches. There is an urgent need for a “health in all policies” framework that synergizes targeted maternal-neonatal interventions (e.g., risk-based IAP and vaccination development) with broad investments in infrastructure, education, and poverty alleviation. Only by synchronizing strengthened healthcare with socioeconomic advancement can we break the stagnation in high-burden regions and ensure that no neonates are left behind.

## Data Availability

All relevant data are within the manuscript and its Supporting Information files.

## Acknowledgments

The authors acknowledge the Global Burden of Disease Study 2021 collaborators for making the data publicly available. We would like to thank Editage (www.editage.cn) for English language editing.

## References

[1] Patras KA, Nizet V. Group B streptococcal maternal colonization and neonatal disease: molecular mechanisms and preventative approaches. Front Pediatr 2018;6:27. doi:10.3389/fped.2018.00027.

[2] Madrid L, Seale AC, Kohli-Lynch M, Edmond KM, Lawn JE, Heath PT, et al. Infant Group B streptococcal disease incidence and serotypes worldwide: systematic review and meta-analyses. Clin Infect Dis 2017;65(Suppl 2):S160–72. doi:10.1093/cid/cix656.

[3] Gonçalves BP, Procter SR, Clifford S, Koukounari A, Paul P, Lewin A, et al. Estimation of country-level incidence of early-onset invasive Group B Streptococcus disease in infants using Bayesian methods. PLOS Comput Biol 2021;17:e1009001. doi:10.1371/journal.pcbi.1009001.

[4] Gonçalves BP, Procter SR, Paul P, Chandna J, Lewin A, Seedat F, et al. Group B streptococcus infection during pregnancy and infancy: estimates of regional and global burden. Lancet Glob Health 2022;10:e807–19. doi:10.1016/S2214-109X(22)00093-6.

[5] GBD 2021 Diseases and Injuries Collaborators. Global incidence, prevalence, years lived with disability (YLDs), disability-adjusted life-years (DALYs), and healthy life expectancy (HALE) for 371 diseases and injuries in 204 countries and territories and 811 subnational locations, 1990–2021: a systematic analysis for the Global Burden of Disease Study 2021. Lancet 2024;403:2133–61. doi:10.1016/S0140-6736(24)00757-8.

[6] Pangerl S, Sundin D, Geraghty S. Group B streptococcus screening guidelines in pregnancy: A critical review of compliance. Matern Child Health J 2021;25:257–67. doi:10.1007/s10995-020-03113-z.

[7] Zaka N, Umar M, Ahmad AM, Ahmad I, Reza TE, Sarfraz M, et al. Equity trends for the UHC service coverage sub-index for reproductive, maternal, newborn and child health in Pakistan: evidence from demographic health surveys. Int J Equity Health 2023;22:230. doi:10.1186/s12939-023-02043-w.

[8] Stevens GA, Alkema L, Black RE, Boerma JT, Collins GS, Ezzati M, et al. Guidelines for Accurate and Transparent Health Estimates Reporting: the GATHER statement. Lancet 2016;388:e19–e23. doi:10.1016/S0140-6736(16)30388-9.

[9] Hosseinpoor AR, Bergen N, Schlotheuber A. Promoting health equity: WHO health inequality monitoring at global and national levels. Glob Health Action 2015;8:29034. doi:10.3402/gha.v8.29034.

[10] Moreno-Betancur M, Latouche A, Menvielle G, Kunst AE, Rey G. Relative index of inequality and slope index of inequality: a structured regression framework for estimation. Epidemiology 2015;26:518–27. doi:10.1097/EDE.0000000000000311.

[11] Schrag S, Gorwitz R, Fultz-Butts K, Schuchat A. Prevention of perinatal group B streptococcal disease. Revised guidelines from CDC. MMWR Recomm Rep 2002;51:1–22.

[12] Colbourn T, Gilbert R. An overview of the natural history of early onset group B streptococcal disease in the UK. Early Hum Dev 2007;83:149–56. doi:10.1016/j.earlhumdev.2007.01.004.

[13] Trotter CL, Alderson M, Dangor Z, Ip M, Le Doare K, Nakabembe E, et al. Vaccine value profile for Group B streptococcus. Vaccine 2023;41(Suppl 2):S41–52. doi:10.1016/j.vaccine.2023.04.024.

[14] Lozano R, Naghavi M, Foreman K, Lim S, Shibuya K, Aboyans V, et al. Global and regional mortality from 235 causes of death for 20 age groups in 1990 and 2010: a systematic analysis for the Global Burden of Disease Study 2010. Lancet 2012;380:2095–128. doi:10.1016/S0140-6736(12)61728-0.

[15] O’Sullivan CP, Lamagni T, Patel D, Efstratiou A, Cunney R, Meehan M, et al. Group B streptococcal disease in UK and Irish infants younger than 90 days, 2014–15: a prospective surveillance study. Lancet Infect Dis 2019;19:83–90. doi:10.1016/S1473-3099(18)30555-3.

[16] Van Kassel MN, Gonçalves BP, Snoek L, Sørensen HT, Bijlsma MW, Lawn JE, et al. Sex differences in long-term outcomes after Group B streptococcal infections during infancy in Denmark and the Netherlands: national cohort studies of neurodevelopmental impairments and mortality. Clin Infect Dis 2022;74(Suppl 1):S54–63. doi:10.1093/cid/ciab822.

[17] Fish EN. The X-files in immunity: sex-based differences predispose immune responses. Nat Rev Immunol 2008;8:737–44. doi:10.1038/nri2394.

[18] Oliveira LMA, Prasad N, Lynfield R, Ip M, Sanou S, Neves FPG, et al. WHO defeating meningitis symposium, 3rd international symposium on Streptococcus agalactiae disease (ISSAD) in Rio de Janeiro, Brazil: state-of-the-art overview of S. agalactiae meningitis. Vaccine 2025;52:126895. doi:10.1016/j.vaccine.2025.126895.

[19] Shibata M, Matsubara K, Matsunami K, Miyairi I, Kasai M, Kai M, et al. Epidemiology of group B streptococcal disease in infants younger than 1 year in Japan: a nationwide surveillance study 2016–2020. Eur J Clin Microbiol Infect Dis 2022;41:559–71. doi:10.1007/s10096-021-04396-y.

[20] Tazi A, Disson O, Bellais S, Bouaboud A, Dmytruk N, Dramsi S, et al. The surface protein HvgA mediates group B streptococcus hypervirulence and meningeal tropism in neonates. J Exp Med 2010;207:2313–22. doi:10.1084/jem.20092594.

[21] Russell NJ, Seale AC, O’Driscoll M, O’Sullivan C, Bianchi-Jassir F, Gonzalez-Guarin J, et al. Maternal colonization with Group B streptococcus and serotype distribution worldwide: systematic review and meta-analyses. Clin Infect Dis 2017;65(Suppl 2):S100–11. doi:10.1093/cid/cix658.

[22] GBD 2021 Causes of Death Collaborators. Global burden of 288 causes of death and life expectancy decomposition in 204 countries and territories and 811 subnational locations, 1990–2021: a systematic analysis for the Global Burden of Disease Study 2021. Lancet 2024;403:2100–32. doi:10.1016/S0140-6736(24)00367-2.

